# Relations of parameters for describing the epidemic of COVID―19 by the Kermack―McKendrick model

**DOI:** 10.1101/2020.02.26.20027797

**Authors:** Toshihisa Tomie

## Abstract

In order to quantitatively characterize the epidemic of COVID-19, useful relations among parameters describing an epidemic in general are derived based on the Kermack–McKendrick model. The first relation is **1/*****τ***_***grow***_ =**1/*****τ***_***trans***_−***1/******τ***_***inf***_, where ***τ***_*grow*_ is the time constant of the exponential growth of an epidemic, ***τ***_*trans*_ is the time for a pathogen to be transmitted from one patient to uninfected person, and the infectious time ***τ***_*inf*_ is the time during which the pathogen keeps its power of transmission. The second relation ***p(***∞) ≈ **1**−**exp(**−**(*R***_***0***_−**1)/0**.**60)** is the relation between *p(*∞), the final size of the disaster defined by the ratio of the total infected people to the population of the society, and the basic reproduction number, *R*_*0*_, which is the number of persons infected by the transmission of the pathogen from one infected person during the infectious time. The third relation **1/*****τ***_***end***_ = **1/*****τ***_***inf***_−**(1**−***p(***∞**))/*****τ***_***trans***_ gives the decay time constant ***τ***_*end*_ at the ending stage of the epidemic. Derived relations are applied to influenza in Japan in 2019 for characterizing the epidemic.

## 1. Introduction

We reported the understanding of the present status and forecasting of pneumonia by COVID―19 in China which is supposed to have originated in Wuhan by analyzing the data up to February 11 (ref.1). In ref.1, we clarified that the behavior of the epidemic was different in different regions and that the outbreak was well described by a Gaussian as was for influenza in Japan. We reported the following; 1. the epidemic in China passed the peak in the beginning of February, 2. the date of the epidemic peak was different by region and was the latest in Wuhan. For more than 10 days after our forecast, the epidemic closely followed our forecast. The new patients outside Wuhan decreased to less than 4 % of that at the peak around February 5.

Although the epidemic in China is near the end, patients by COVID―19 are found in many other countries, and COVID―19 is still a big fear of the people over the world. In order to forecast the epidemic of COVID―19 in other countries, we need to theoretically characterize the COVID―19 epidemic by fitting a model calculation to the data observed in China. We choose the Kermack― McKendrick model (ref.2) for our analysis. The Kermack―McKendrick model was proposed as early as 1927, but still, it is the basis of many modified models for describing epidemics.

## 2. Present status of COVID―19 in China

Figure 1 shows the changes of newly infected people of COVID―19 in China. The data were taken from refs. 3 and 4. Up to 40^th^ day, *i*.*e*., February 9, the outbreak of the epidemic was described well by a Gaussian as was clarified in ref.1. The fitting Gaussians are 850*exp(− ((x− 33)/8)^2^) for mainland China except Hubei province, 1100*exp(− ((x − 34.2)/7)^2^) for Hubei province except Wuhan, and 1850*exp(− ((x − 38)/7)^2^) for Wuhan. In ref.1, we mentioned that influenza in Japan shifted from a Gaussian to an exponential decay at the skirt of the epidemic. The decay of COVID―19 in China was forecasted by assuming an exponential decay. Reference 1 was written on February 12. Since that day, the epidemic of COVID―19 in China has been changing as forecasted. On February 20, the number of newly infected people was only one-thirtieth of that at the peak in mainland China except Hubei province. The fitted exponential decay curves are 350*exp(− (x−42)/4.5) for mainland China except Hubei province, 800*exp(−(x−40)/5) for Hubei province except Wuhan, and 2700*exp(−(x−40)/5) for Wuhan.

**FIG.1;.**
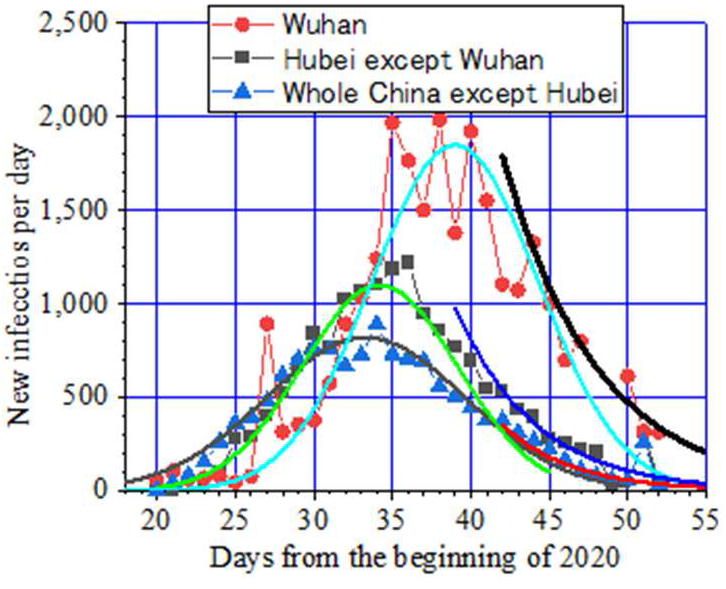
Present status of COVID―19 in China. The shapes of the epidemic in three regions changed from a Gaussian to an exponential decay around February 10.

We want to analyze the epidemic of COVID―19 in detail by applying a model. As a preparation of the analysis, useful relations of parameters describing an epidemic are derived in the present paper.

## 3. Derivation of the relations among parameters

Useful relations of parameters of an epidemic are Eqs. (7) to (10) in the following, which are derived from the Kermack―McKendrick model (ref.2). The number of susceptible people in the group is set as *S*_*0*_, the number of infected persons is *I(t)*, the number of persons who have been infected and recovered to obtain immunity or have died is *R(t)*, the number of persons who is susceptible but not yet infected is *S(t). S*_*0*_ *= S(t) + I(t) + R(t)*. When the transmission power of the disease is set as *β* and the recovery rate from the infection is *γ*, the epidemic of the disease is given by the following three differential equations.

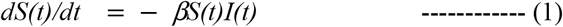

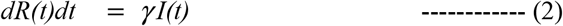

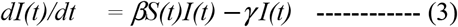

We define the initial transmission time constant, ***τ***_*trans*_, after which a pathogen is transmitted to one susceptible host in the early stage of the epidemic and the infectious time of a pathogen, ***τ***_*inf*_, after which the pathogen loses the power of transmission as follows,

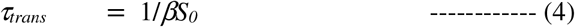

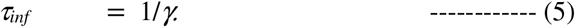

The basic reproduction number, *R*_*0*_, which is the number of persons infected by the transmission of a pathogen from one infected person during the infectious time, is given by

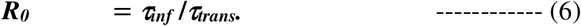

In the following, we see that an epidemic starts with exponential growth with a time constant ***τ***_*grow*_ given by

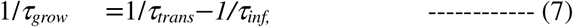

and decays exponentially with a time constant ***τ***_*end*_ given by

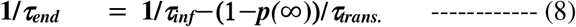

Here, *p(*∞) is the final size of the disaster which is defined by the ratio of the total number of the infected people, *R(*∞), to the population of the society, *S*_*0*._ As shown later, *p(*∞) is approximated as,

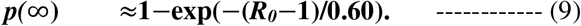

By using *R*_*0*_, Eq. (7) is rewritten as

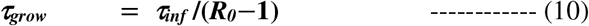

*R*_*0*_, ***τ***_*inf*,_ and *p(*∞) are parameters describing an epidemic. The above relations, Eqs. (7) to (10), are derived as follows.

In the beginning of the epidemic, *I(t)* and *R(t)* are negligible compared to *S*_*0*_, and Eq.(3) becomes

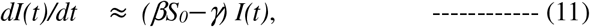

and we get

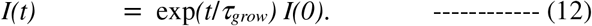

Thus, Eq. (7) is derived.

The condition of spreading the disease is ***τ***_*grow*_> 0, and then, *R*_*0*_ > 1. For this condition, *S*_*0*_ should be larger than the transmission threshold *S*_*th*_ given by

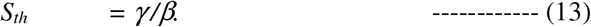

As infection proceeds, *S(t)* reduces. When *βS(t)-γ* becomes negative, the number of infected persons *I(t)* begins to decrease according to Eq. (3), and eventually, *I*(*t*) becomes 0. When *I (t)* is 0, the reduction of the number of the uninfected susceptible persons *S(t)* stops, as seen in Eq. (1), and *S(t)* converges to *S(*∞).

By dividing Eq. (3) by Eq. (1), we get

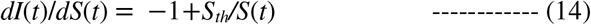

and then we get

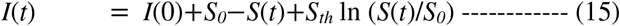

Because *I(*∞*)=0* and *I*(0) << *S*_*0*_, we get

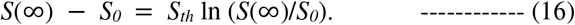

We write the ratio of the population of the recovered person *R* (t) to *S*_*0*_ as *p* (t),

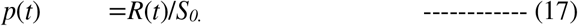

By replacing *S(*∞*)/S*_*0*_ by *p*(∞), Eq. (16) becomes

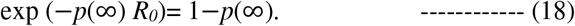

In deriving Eq. (18), we employed the relation

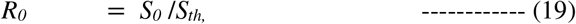

which is derived from Eqs. (4), (5), (6), and (13). The number *p(*∞) is referred as the “final size” and Eq. (18) is referred as the “final size equation”.

At the shrinking stage of the spread of the disease, *S(t)* converges to *S(*∞). Then, Eq. (3) becomes,

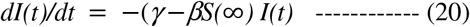

which gives exponential decay of the patients.

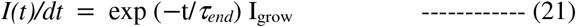

The time constant of the decay ***τ***_*end*_ is given by

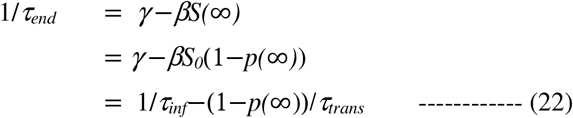

Thus, Eq. (8) is derived.

The final size equation was numerically solved and the result is shown by the solid curve in Fig.2. As *R*_*0*_ increases from 1, *p (*∞) increases from zero and saturates at a large *R*_*0*._ When *R*_*0*_ is larger than 2.5, more than 90 % of people in the group are infected. As shown by the dotted curve in Fig.2, *p*(∞) can be approximated by the equation, 1-exp(−(*R*_*0*_−1)/0.60). Thus, Eq, (9) is derived.

**FIG.2:**
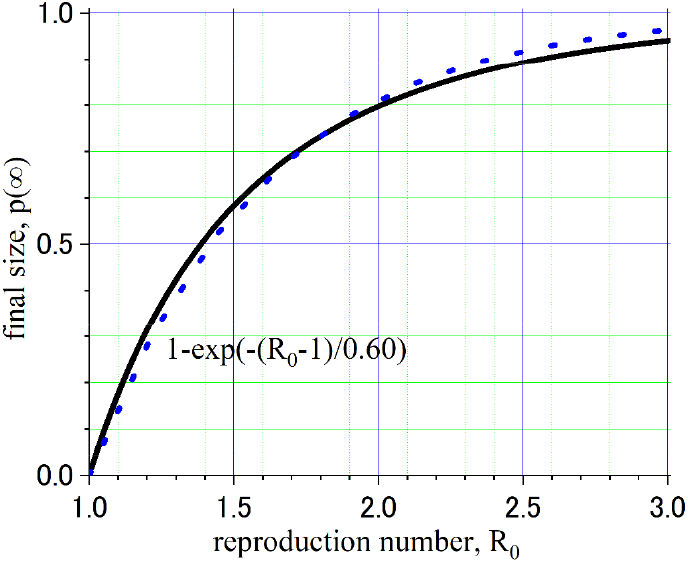
Numerically calculated final size *p(∞)* of an epidemic as a function of re-production number, *R*_*0*_.

## 4. Applying the model to the epidemic of influenza in Japan

We apply the above model to the epidemic of influenza in Japan as cited in ref.1 as a reference for the general epidemic. Figure 3 shows the influenza epidemic in Japan over the past decade (ref.5). We choose influenza in 2019, which is referred as JpnInf2019 in this paper. In ref.1, we fitted a Gaussian to JpnInf2019. As shown by the red solid curve in Fig. 4, the model described above reproduces JpnInf2019 better than a simple Gaussian. The parameters were ***τ***_*tran s*_= 0.52 weeks and ***τ***_*inf*_ = 1 week.

**FIG.3:**
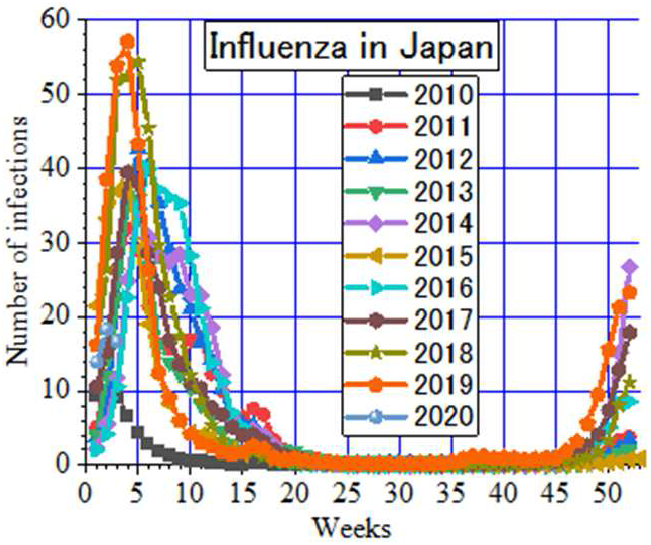
Epidemic of infuenza in Japan over the past decade.

**FIG.4:**
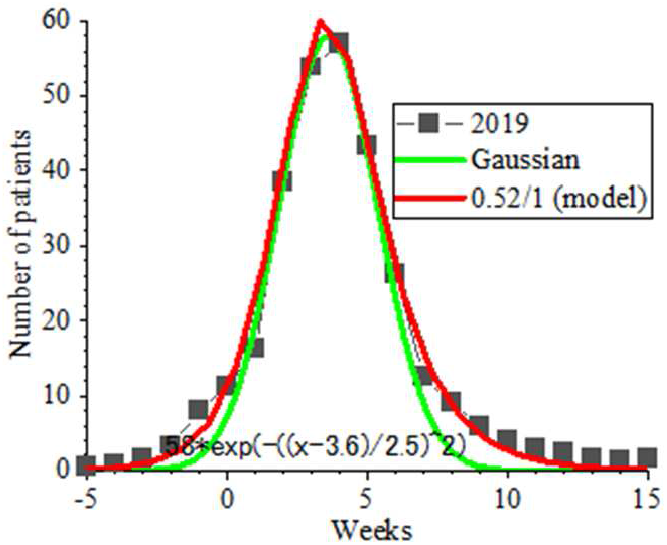
Reproducing JpnInf2019 by the model with ***τ***_*trans*_= 0.52 weeks and ***τ***_*inf*_ = 1 week.

From the values of ***τ***_*trans*_ and ***τ***_*inf*_ for fitting, the reproduction number *R*_*0*_ = 1.92 and the final size estimated by Eq. (18) is *p*(∞) = 0.77. Ministry of Health, Labor and Welfare of the Japanese government reported that the estimated number of patients who consulted doctors was about 12,000, 000 over the country (ref. 6). This estimated value was only 10 % of the population of Japan and far smaller than *p*(∞) = 77 %. There are three possibilities for the reason. The first is that most patients did not go to a hospital and the real number of patients was 92,400, 000. The second is that only 1/8 of the area in Japan was the reach of the pathogen. The third is a combination of the first and the second reasons. Thus, we realize the evaluation of the final size *p*(∞) is difficult.

While reproduction of the epidemic by the model is fairly good near the peak of the epidemic, but at the far skirt, deviation of the data from the model is not negligible as seen in Fig.5 which shows a log plot of Fig.4.In the model fitting, the time constant was 1.15 weeks at the rising and the decay time constant was 1,7 weeks. Hence, ***τ***_*grow*_ calculated by Eq. (7) was 1.1 weeks. But in JpnInf2019, the reported number of patients increased exponentially with a time constant of 1.5 weeks and decreased exponentially at the ending stage with a time constant of 2.5 weeks. By increasing ***τ***_*trans*_ to 0.61 weeks from 0.52 weeks, the time constant of the rising of the epidemic can be increased to 1.5 weeks in the model calculation, but the width of the epidemic in the model calculation was too wider than the real one as shown in Fig.6.

**FIG.5:**
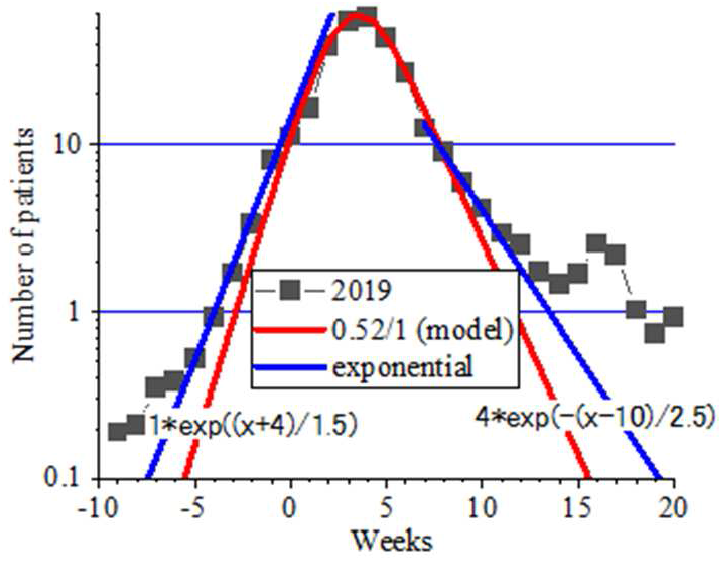
A log plot of Fig.4. At the skirt, the increase and the decrease of the epidemic is slower than the model.

**FIG.6:**
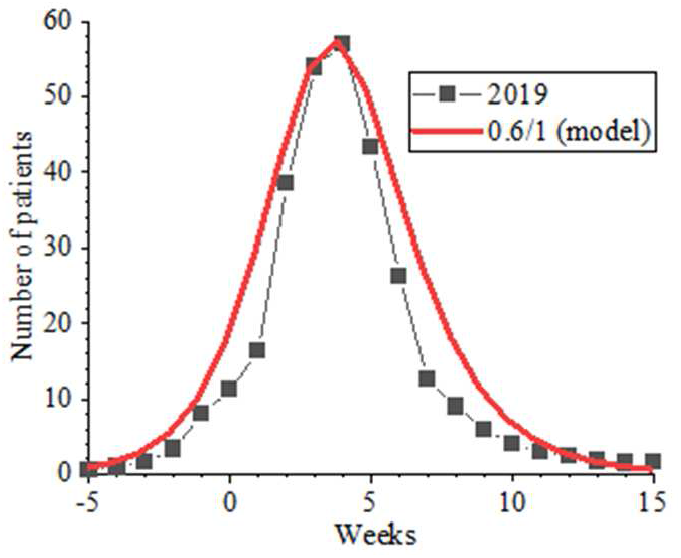
If the observed ***τ***_***grow***_ and ***τ***_*end*_ are used, the model cannot reproduce the main part of the epidemic.

In all the cases of the epidemic, there should be a difference between the reported number of patients and the real number of infected people. Not all infected people will go to a hospital and not all patients are not inspected by medical institutes. In analyzing statistics, we assume the ratio of the reported number to the real number is constant during the epidemic, but often the assumption can be wrong. We expect the ratio is smaller in the beginning and at the ending of an epidemic. Then, the time constant of the “apparent rise” and “apparent ending” will be shorter than the real ones. However, this expectation was the opposite in Fig.5.

The slower increase and the slower decrease at the skirt of the epidemic could suggest the transmission power *β* of the virus may change in time, which, we think, is not plausible. At present, we do not know the reason why the simple Kermack–McKendrick cannot reproduce the whole epidemic including the skirt parts. Thus, from the analysis of JpnInf2019 by the model described above, we learn that it is important to remember that the time constant of the “apparent” increase of the epidemic in the early stage does not re-produce the whole epidemic.

The above information helps greatly to understand the to-be-planned analysis of COVID―19.

## 5. Summary

Useful relations of parameters of an epidemic are derived by following the Kermack–McKendrick model. The first relation is 1/***τ***_*grow*_ = 1/***τ***_*trans*_−*1/**τ***_*inf*_. The final size of the disaster was numerically calculated and we found it is given by *p(*∞) ≈ 1−exp(−(*R*_*0*_ −1)/0.60). The third relation 1/***τ***_*end*_ = 1/***τ***_*inf*_ −(1−*p(*∞))/***τ***_*trans*_ gives the decay time constant ***τ***_*end*_ at the last stage of the epidemic.

By applying the model, we found that the epidemic of influenza in Japan in 2019 was re-produced by the parameters;***τ***_*trans*_ = 0.52 week and ***τ***_*inf*_ = 1 week and that ***τ***_*grow*_ observed in the early stage can be different from ***τ***_*grow*_ for re-producing the overall epidemic.

## Data Availability

All data referred to in the manuscript is available at references.

